# A Novel Use of a Tissue Oxygenation Monitor at Time of Uterus Transplantation and Hysterectomy – A Feasibility Study

**DOI:** 10.1101/2022.11.09.22280210

**Authors:** Jeremy Applebaum, Dan Zhao, Dan Barry, Nawar Latif, Kathleen O’Neill

**Affiliations:** Department of Obstetrics and Gynecology, University of Pennsylvania Health System, Philadelphia, PA; ViOptix, Inc., Fremont, CA

## Abstract

While thrombosis is the most common indication for graft failure and immediate hysterectomy of a transplanted uterus, there is no optimal method to monitor graft perfusion. In this feasibility study, a near-infrared spectroscopy probe that monitors local tissue oxygenation (StO_2_) was attached to four uterine cervices and three donor cervices at the time of hysterectomy and transplantation respectively. Real-time StO_2_ measurements were gathered at key surgical steps of the hysterectomies and uterus transplantations. The average StO_2_ for the sequential steps of a hysterectomy of baseline, ovarian vessel ligation, contralateral ovarian vessel ligation, uterine vessel ligation, contralateral uterine vessel ligation, and colpotomy was 70.2%, 56.7%, 62.1%, 50.5%, 35.8%, and 8.5% respectively. The average StO_2_ for the sequential steps of uterus transplantation of iliac vein anastomosis, iliac artery anastomosis, contralateral iliac vein anastomosis, contralateral iliac artery anastomosis, and vaginal anastomosis was 8.9%, 27.9%, 56.9%, 65.9%, and 65.2% respectively. As uterine blood supply decreases in a hysterectomy, the measured StO_2_ also decreases, and vice versa for uterus transplantation. Tissue oximetry may be a reliable, non-invasive means of monitoring perfusion of a uterine graft. Additional studies are needed to determine if these devices complement current assessments of uterine graft viability and salvage thrombosed grafts.

## Introduction

Uterus transplantation (UTx) is an emerging intervention that provides women with absolute uterine factor infertility the opportunity to experience pregnancy [1,2]. Stable vascular perfusion is critical for the viability of the uterine allograft, as approximately 20 percent of UTx result in emergent postoperative hysterectomy for thrombosis or graft hypoperfusion [3–5]. Poor UTx perfusion may relate to the complexity of donor-recipient vascular anastomoses and underlying donor atherosclerosis [3,4,6]. Development and application of technologies to accurately detect and prompt intervention on poorly perfused grafts is crucial for ongoing UTx development.

Current methods for postoperative monitoring of graft perfusion include transabdominal or transvaginal doppler ultrasonography or the use of a Cook-Swartz doppler flow probe. The Cook-Swartz probe contains a 20-megahertz crystal ultrasound transducer that is attached to a silicon cuff which is secured around arterial supply to the uterus, connected to stress retention tabs and finally attached to a terminal connector external to the patient. While these strategies have been used to monitor blood flow postoperatively following reconstructive vascular surgery, there are a number of limitations to these diagnostic tools [3,4]. Doppler ultrasonography is limited by documented false-negative findings in thrombosed grafts [4]. The Cook-Swartz probe is also limited in that it is invasive and cannot detect venous thrombosis [3,4]. Angiography can confirm suspected thrombosis after it has already occurred, though this often does not allow for salvageability [4,7]. Angiography also imposes a radiographic contrast load on patients at risk for acute kidney injury given their recent induction with nephrotoxic immunosuppressants [8]. Importantly, these methodologies are all measures of peripheral blood flow and oxygenation (SpO_2_) which may not accurately reflect local tissue oxygenation (StO_2_) due to differences in oxygen delivery and utilization [9].

Near infrared spectroscopy (NIRS) probes may provide noninvasive, real-time monitoring of UTx cervical StO_2_. NIRS probes have improved thrombosis detection and graft salvage rates in breast, extremity, truncal, and head and neck free flaps in plastic surgery literature [10–14]. The objective of this study is to assess feasibility of uterine cervical intra- and postoperative StO_2_ monitoring with a NIRS probe device during UTx.

## Methods

The device being studied is the ViOptix T. Ox Tissue Oximeter (ViOptix, Inc., Fremont, California) and is patent-protected, Food and Drug Administration-approved for tissue oxygenation monitoring. It consists of a 5 mm by 5 mm flat sensor with two NIRS lasers that penetrate tissue up to 1 cm deep and four photoelectric diodes that detect reflected light (Figure 1). Measurements of infrared light scattering and absorption due to local tissue hemoglobin and deoxyhemoglobin concentrations are used to calculate StO_2_. A fiberoptic cable connects to the sensor to a console that displays continuously recorded StO_2_ measurements (Figure 2) [10,11]. The console also displays an indicator of signal quality, a measure of the consistency of StO_2_ readings between the four diodes. A signal quality of at least 80 percent is considered satisfactory for reliability of StO_2_ readings for clinical purposes. While the ViOptix console allows for dual monitoring of two probes, only one channel was used in this study [15].

**Fig. 1.**
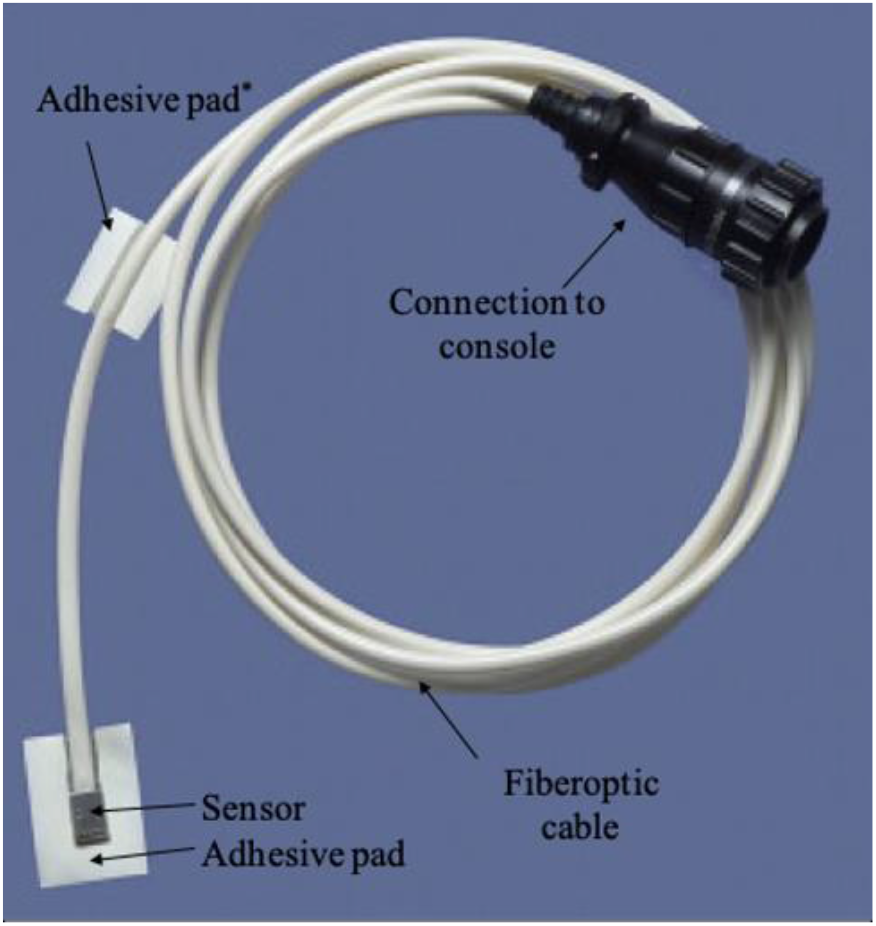
ViOptix probe with adhesive pad attached to fiberoptic cable. *Not used in this study.

**Fig. 2.**
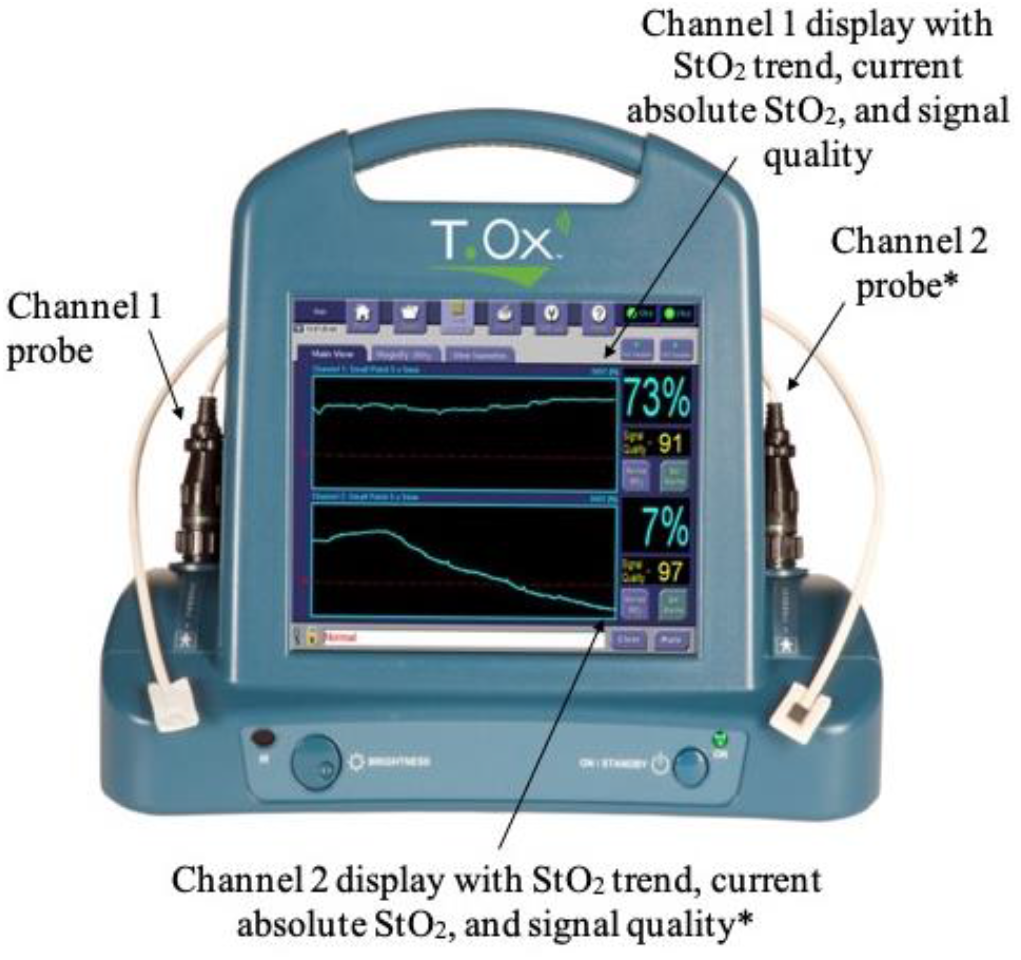
ViOptix console alongside two channel probes with sample display of StO_2_ measurements. *Not used in this study.

As proof of concept, the ViOptix device was used to assess cervical StO2 measurements during hysterectomy. Patients undergoing a total abdominal hysterectomy and bilateral salpingo-oophorectomy (TAH-BSO) with a gynecologic oncologist were enrolled from August 2018 to February 2019. At the beginning of the surgery, vaginal retractors were placed and the ViOptix device was sutured to the cervix. The cable was passed through the vagina and connected to the console. StO_2_ measurements at key surgery steps of the TAH-BSO (ovarian vessel ligation, contralateral ovarian vessel ligation, uterine vessel ligation, contralateral uterine vessel ligation, and colpotomy) were recorded. Once the specimen was freed, the sensor was removed from the cervix and the cable removed through the vagina.

Following the pilot study, the ViOptix device was used for research purposes in subjects undergoing UTx. All patients who underwent UTx at the Hospital of the University of Pennsylvania (Penn Medicine) were enrolled from November 2018 to February 2020. Patients were recruited through Penn Medicine’s Uterine Transplantation for Uterine Factor Infertility (UNTIL) trial.

The ViOptix device was used for both intraoperative and postoperative StO_2_ monitoring in UTx. During backbench preparation of the donor uterine grafts, the ViOptix sensor was sutured to the left lateral aspect of the donor cervix. Once the recipient’s pelvic vasculature was surgically exposed and colpotomy was made, the graft was brought to the operative field and the ViOptix cable was fed through the colpotomy and connected to an external console. StO_2_ measurements were then continuously recorded at five second intervals for the duration of surgery. StO_2_ measurements at key surgical steps of the UTx (donor internal iliac vein anastomosis to recipient external iliac vein, donor internal iliac artery anastomosis to recipient external iliac artery, contralateral donor internal iliac vein anastomosis to recipient external iliac vein, contralateral donor internal iliac artery anastomosis to recipient external iliac artery, and donor and recipient vagina anastomosis) were recorded. Cervical StO_2_ measurements were also recorded for upwards of three days postoperatively. The ViOptix device was removed during scheduled post-UTx exams under anesthesia.

Criteria studied in both the UTx and TAH-BSO patients were the absolute StO_2_ value, the amount of its change (ΔStO_2_) and the rate of its change (ΔStO_2_/Δtime). Given that there is no published use of the ViOptix device in gynecologic surgery, surgical reexploration of a UTx was not performed in this study due to isolated StO_2_ values or StO_2_ trends.

The Institutional Review Board of the University of Pennsylvania gave ethical approval of this work (UNTIL #827853, ViOptix #829582).

## Results

### Participant demographics

Four patients undergoing a TAH-BSO were enrolled. All TAH-BSO patients underwent surgery for suspected gynecologic malignancy, with pathology confirmed malignancy in all but one patient. Only one patient (H2) underwent neoadjuvant chemotherapy.

All three patients who underwent a UTx at Penn Medicine were enrolled. All UTx patients had congenital absence of the uterus. TAH-BSO and UTx participant demographic features are listed in Table 1. To protect potentially personally identifiable information, an age range, rather than precise ages, are presented. Table 1 also displays the pregnancy outcomes of Penn Medicine’s UNTIL trial.

**Table 1.** Uterus transplantation and hysterectomy participant demographics

| UTx | Patient Identifier | Age at surgery (y) | BMI (kg/m<sup>2</sup>) | Race | Donor allograft status | Subsequent pregnancy outcomes |
| --- | --- | --- | --- | --- | --- | --- |
|  | T1 | 30-35 | 19 | White | Deceased donor in her 20s, three prior vaginal deliveries | 1 live-born infant, cesarean hysterectomy |
|  | T2 | 25-30 | 32 | White | Deceased donor in her 30s, two prior vaginal deliveries | 2 live-born infants: cesarean section, followed by cesarean hysterectomy |
|  | T3 | 30-35 | 21 | White | Living donor in her 40s, one prior vaginal and one cesarean section | 2 live-born infants: cesarean section, followed by cesarean hysterectomy |
| TAH-BSO | Patient Identifier | Age at surgery (y) | BMI (kg/m<sup>2</sup>) | Race | Pathology |  |
|  | H1 | 60-65 | 28 | Black | Uterine carcinosarcoma |  |
|  | H2 | 60-65 | 42 | White | Ovarian serous carcinoma |  |
|  | H3 | 70-75 | 37 | Black | Ovarian mucinous cystadenoma |  |
|  | H4 | 55-60 | 30 | Black | Endometrial endometrioid carcinoma |  |

### Hysterectomy and bilateral salpingo-oophorectomy intraoperative monitoring

A combined 2,760 unique StO_2_ measurements were gathered for the four TAH-BSO patients over a time range of 1:09:43 – 1:52:14 (hours:minutes:seconds). Figure 3 displays the cervical StO_2_ for each TAH-BSO patient and average by key surgical step. The average cervical StO_2_ for the sequential steps of baseline, ovarian vessel ligation, contralateral ovarian vessel ligation, uterine vessel ligation, contralateral uterine vessel ligation, and colpotomy was 70.2%, 56.7%, 62.1%, 50.5%, 35.8%, and 8.5% respectively.

**Fig. 3.**
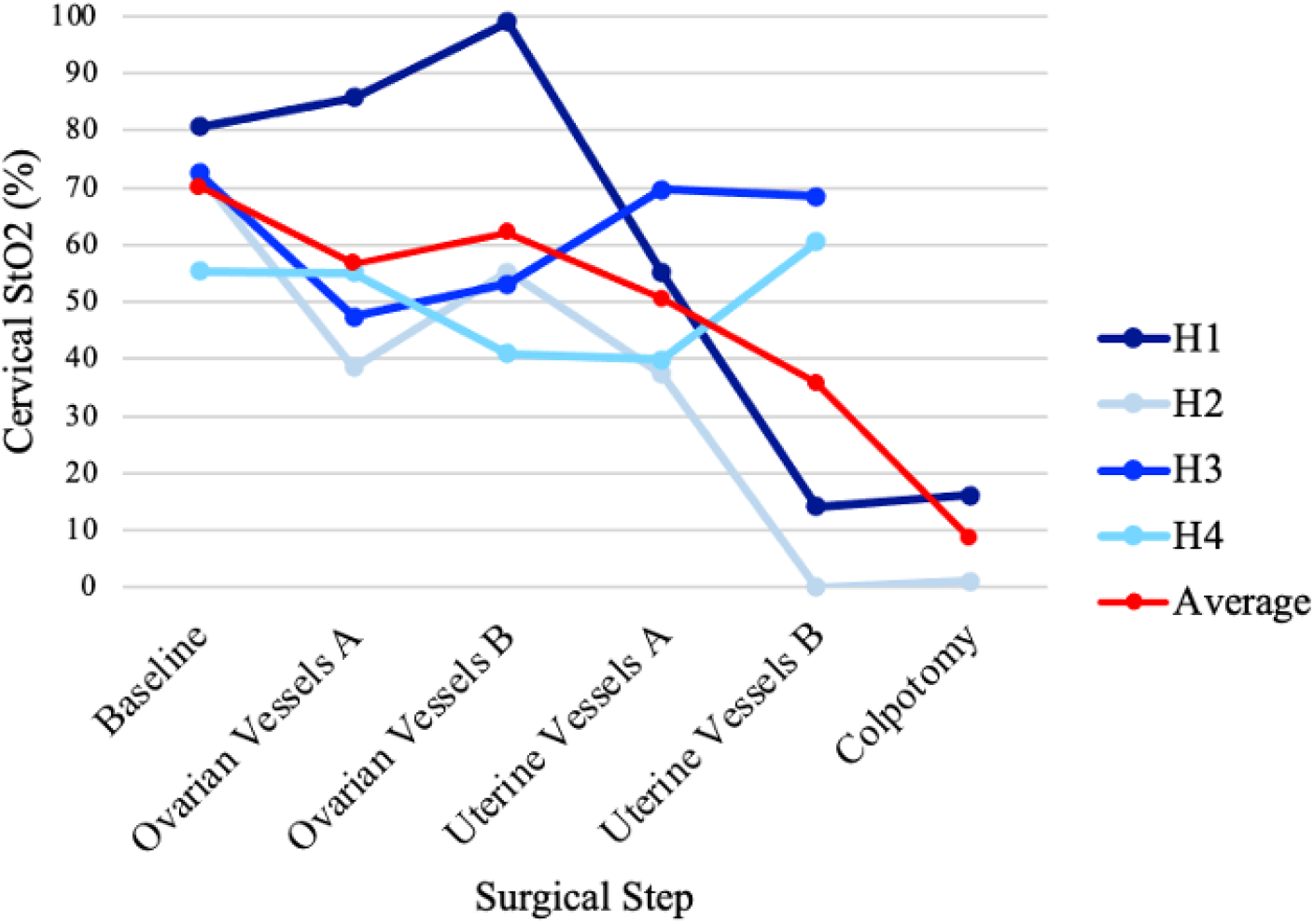
Cervical StO_2_ by key step of hysterectomy and bilateral salpingo-oophorectomy

### Uterus transplantation intraoperative monitoring

Figure 4 displays the cervical StO_2_ for each UTx patient and average by key surgical step. The average cervical StO_2_ for the sequential steps of uterus transplantation of iliac vein anastomosis, iliac artery anastomosis, contralateral iliac vein anastomosis, contralateral iliac artery anastomosis, and vaginal anastomosis was 8.9%, 27.9%, 56.9%, 65.9%, and 65.2% respectively.

**Fig. 4.**
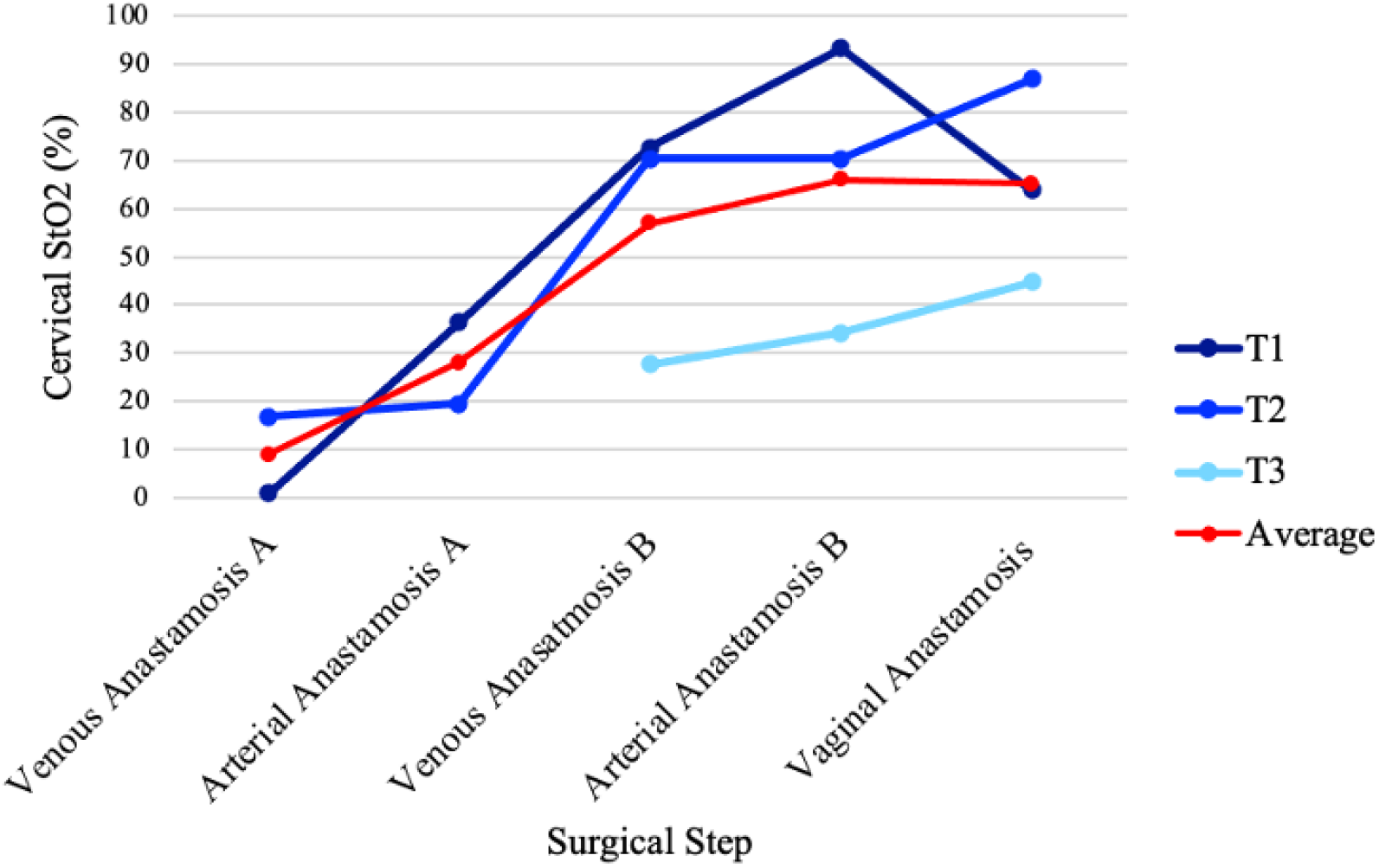
Cervical StO_2_ by key step of uterus transplantation

### Postoperative monitoring of uterus allograft

A combined 90,746 unique StO_2_ measurements were gathered for the three UTx patients over a time range of 0:08:56:34 – 3:04:04:51 (days:hours:minutes:seconds) including both operative time and postoperative monitoring.

Of note, in patient T1, approximately 19 hours postoperatively, an abrupt decline in absolute cervical StO_2_ from 86.3% to 7.7% followed by eventual spontaneous resolution to StO_2_ measurements greater 90% approximately 60 minutes later (Figure 5). Given no clinical decisions were made using ViOptix, no interventions were performed. Routine doppler ultrasonography three hours later demonstrated adequate bilateral uterine arterial flow. On postoperative day (POD) 6, the patient underwent computed tomography of her abdomen and pelvis for an unrelated indication and an incidental occlusion was noted along the right external iliac artery. Magnetic resonance angiography confirmed an occluded graft between recipient right external iliac artery and donor right internal iliac artery with reconstitution at the level of the right common femoral artery via the inferior epigastric artery. Right venous and left arterial and venous vascular patency was noted. Vascular surgery was consulted and given lack of symptoms and persistent radiographic uterine enhancement, likely through collaterals or contralateral supply, decision was made to leave the graft in-situ. The patient was initiated on therapeutic anticoagulation, which was continued for 8 weeks postoperatively.

**Figure 5:**
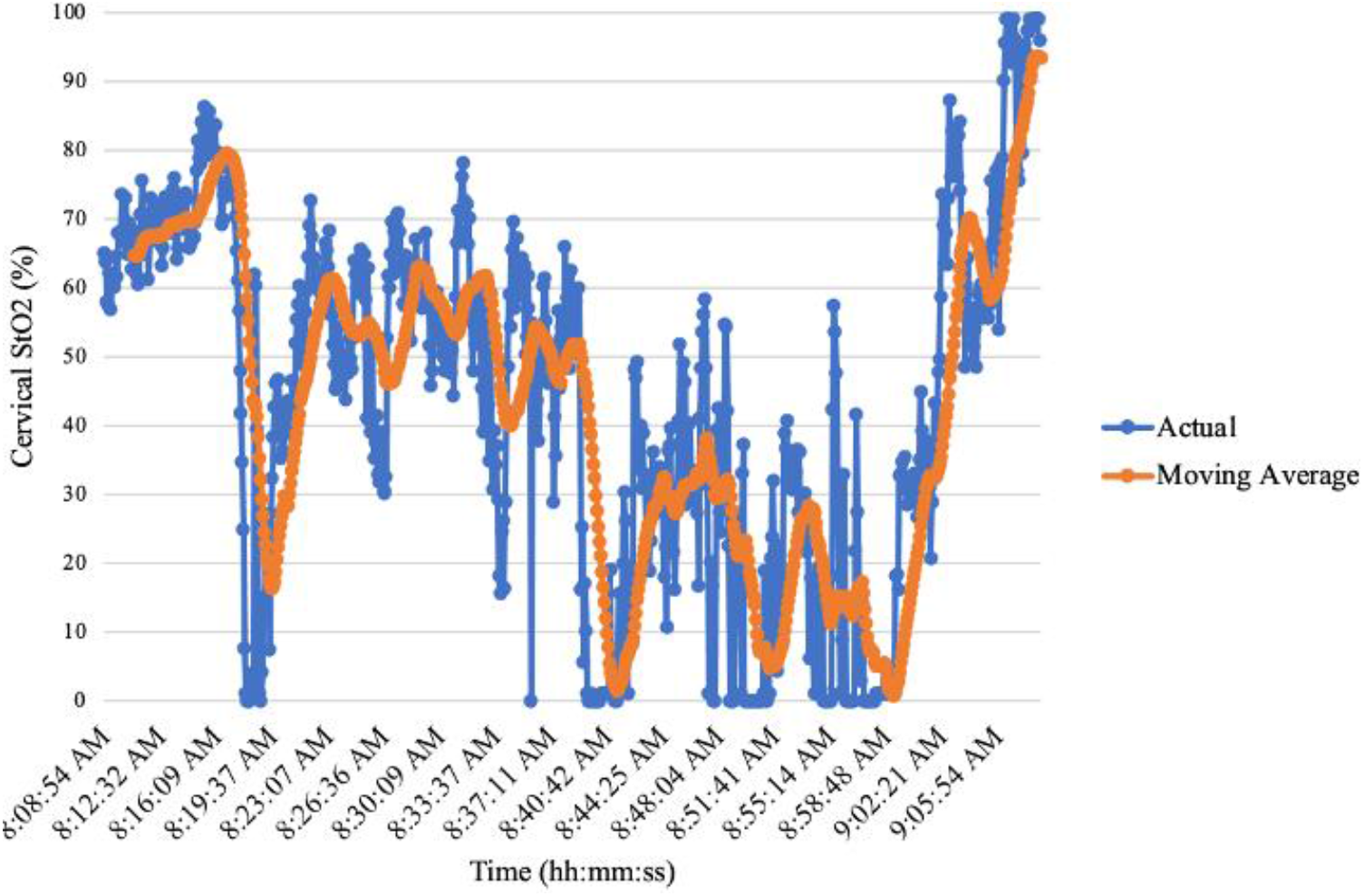
Abrupt decline in patient T1 cervical StO_2_ on postoperative day 1. Actual values are depicted in blue. A 10 point interval moving average is depicted in orange.

The ViOptix probes were removed on POD3 or 4 during scheduled exams under anesthesia. At the time of removal there was poor application of the ViOptix sensor to the cervix in all three cases.

## Discussion

In this feasibility study, we demonstrate a relative increase and decrease in human cervical StO_2_ throughout the surgical steps of UTx and TAH-BSO respectively. This demonstrates that the ViOptix device is a feasible means of detecting relative StO_2_ trends of the human cervix. Given the novelty of NIRS monitoring in uterus transplantation, there are no established recommendations for graft intervention due to StO_2_ trends alone. While one patient experienced an abrupt postoperative StO_2_ decline and a subsequent incidentally detected graft thrombosis, it is unclear if these events are causally related, particularly in the presence of normal arterial doppler flow and intermittently poor signal quality.

StO_2_ has been described as an adjunct to clinical examination in monitoring graft perfusion. The ViOptix device, and other NIRS-based point probes, have demonstrated efficacy in both detecting and preventing graft thrombosis in various free flap transfers [10–13]. Given that vascular thrombosis is the leading cause of immediate post-UTx hysterectomy, NIRS may have potential application as a non-invasive means of postoperative UTx graft monitoring [3,16].

There is limited data on the application of NIRS probes with surgery of the human uterus and cervix. Prior work has used NIRS probes to demonstrate that different optical properties of the human cervix occur throughout the menstrual cycle, pregnancy trimesters, during obstetric cervical ripening, and in the presence of dysplasia [17–20]. Uterine corpus StO_2_ mapping as measured through a spectroscopy monitor affixed to a laparoscope has also been performed on UTx in sheep and rabbit models [21].

There are several potential advantages of NIRS probes for UTx graft monitoring. NIRS probes provide real-time, continuous monitoring unlike intermittent ultrasonographic or clinical assessments. This may allow for decreased resource allocation for postoperative graft care [10]. These devices are also non-invasive and can be both affixed and removed from the cervix through simple speculum examination. NIRS probes also provide information on oxygenation in the tissue of interest as opposed to its vascular supply, which may differ due to oxygen dissociation from local temperature or pH [9,22].

This study is not without limitations. The purpose of this study was to assess feasibility of the application of the ViOptix device in small pilot sample, and we are unable to study whether ViOptix detects graft thrombosis and improves salvage rates. Given that NIRS probes are commonly single-use, cost may be an additional limitation to routine application [10]. However, given the costs associated with reoperations and graft loss, further studies are necessary to ascertain cost-effectiveness of this new technology with UTx. Though a downtrend in cervical StO_2_ was noted with the subsequent steps of a hysterectomy, most of these patients had a gynecologic malignancy, which may result in aberrant cervical perfusion either due to the malignancy itself or antineoplastic treatments. While cervical application allows for NIRS probes to be non-invasively applied, it is unclear if cervical StO_2_ is an adequate proxy for entire uterine perfusion, especially considering a limited depth of laser penetration. Lastly, there may be issues with full contact of the flat, paddle shaped ViOptix probe on a naturally curved cervix, particularly given postoperative changes to the cervical contour due to edema, inflammation, or ischemia-reperfusion effects.

More important than the development of novel technology to monitor graft perfusion, however, is the establishment of criteria to prompt clinical intervention. Identification of isolated StO_2_ levels or a decreasing StO_2_ trend that may prompt more intensive monitoring or surgical intervention is needed. Correlates from breast, head and neck, and truncal free flap literature demonstrates that StO_2_ values between 30-40% and a StO_2_ decrease of 15-20% within one hour is predictive of flap failure [11,12,23]. Larger scale studies are needed to identify StO_2_ criteria through NIRS probes that predict UTx failure.

While we have demonstrated the feasibility of a NIRS probe for intra- and postoperative StO_2_ monitoring during UTx, additional larger-scale studies are needed to determine whether these devices may improve detection of thrombotic graft compromise and improve salvage rates.

## Conclusion

In conclusion, the ViOptix NIRS probe applied to cervices at the time of hysterectomy and uterine transplantation demonstrated a relative decrease and increase in StO_2_ respectively. Development and refinement of technologies, such as the ViOptix device, to non-invasively monitor uterine perfusion may be an important factor in streamlining this emerging therapy for absolute uterine factor infertility.

## Supporting information

ICMJE Disclosure Form

STROBE Checklist

## Data Availability

Data underlying the results presented in this paper may be obtained from the authors upon reasonable request.

## Funding

The ViOptix T. Ox Tissue Oximeters used in this study were provided at no cost by ViOptix, Inc.

## Acknowledgements

N/A

## Disclosures

DB is employed by ViOptix, Inc.

## Notes

### Clinical Trial

NCT03307356

## Reference

1. M. Brännström, L. Johannesson, H. Bokström, N. Kvarnström, J. Mölne, P. Dahm-Kähler, A. Enskog, M. Milenkovic, J. Ekberg, C. Diaz-Garcia, M. Gäbel, A. Hanafy, H. Hagberg, M. Olausson, and L. Nilsson, “Livebirth after uterus transplantation,” Lancet 385, 607–616 (2015).

2. M. Brännström, M. A. Belfort, and J. M. Ayoubi, “Uterus transplantation worldwide: clinical activities and outcomes,” Curr Opin Organ Transplant 26, 616–626 (2021).

3. J. Kristek, L. Johannesson, R. Novotny, D. Kachlik, and J. Fronek, “Human uterine vasculature with respect to uterus transplantation: A comprehensive review,” J. Obstet Gynaecol Res 46, 2199–2220 (2020).

4. G. Testa, E. C. Koon, L. Johannesson, G. J. McKenna, T. Anthony, G. B. Klintmalm, R. T. Gunby, A. M. Warren, J. M. Putman, G. dePrisco, J. M. Mitchell, K. Wallis, and M. Olausson, “Living Donor Uterus Transplantation: A Single Center’s Observations and Lessons Learned From Early Setbacks to Technical Success,” Am J Transplant 17, 2901–2910 (2017).

5. M. Brännström, L. Johannesson, P. Dahm-Kähler, A. Enskog, J. Mölne, N. Kvarnström, C. Diaz-Garcia, A. Hanafy, C. Lundmark, J. Marcickiewicz, M. Gäbel, K. Groth, R. Akouri, S. Eklind, J. Holgersson, A. Tzakis, and M. Olausson, “First clinical uterus transplantation trial: a six-month report,” Fertil Steril 101, 1228–1236 (2014).

6. B. Jones, S. Saso, T. Bracewell-Milnes, M. Thum, J. Nicopoullos, C. Diaz-Garcia, P. Friend, S. Ghaem-Maghami, G. Testa, L. Johannesson, I. Quiroga, J. Yazbek, and J. Smith, “Human uterine transplantation: a review of outcomes from the first 45 cases,” BJOG: Int J Obstet Gy 126, 1310–1319 (2019).

7. H. Leonhardt, A. Thilander-Klang, J. Båth, M. Johannesson, N. Kvarnström, P. Dahm-Kähler, and M. Brännström, “Imaging evaluation of uterine arteries in potential living donors for uterus transplantation: a comparative study of MRA, CTA, and DSA,” Eur Radiol (2021).

8. U. C. Perni, E. Y. Wang, and A. R. Gregg, “Antepartum Care of the Uterus Transplant Patient: The Experience of 3 Successful US Centers,” Clin Obstet Gynecol 65, 84–91 (2022).

9. J. McNulty, M. Born, and R. S. Pozos, “Near-Infrared Spectroscopy (NIRS),” in Springer Handbook of Medical Technology, R. Kramme, K.-P. Hoffmann, and R. S. Pozos, eds. (Springer Berlin Heidelberg, 2011), pp. 423–438.

10. S. J. Lin, M.-D. Nguyen, C. Chen, S. Colakoglu, M. S. Curtis, A. M. Tobias, and B. T. Lee, “Tissue Oximetry Monitoring in Microsurgical Breast Reconstruction Decreases Flap Loss and Improves Rate of Flap Salvage,” Plast Reconst 127, 1080–1085 (2011).

11. M. H. Steele, “Three-Year Experience Using Near Infrared Spectroscopy Tissue Oximetry Monitoring of Free Tissue Transfers,” Ann Plast Surg 66, 540–545 (2011).

12. A. Keller, “A New Diagnostic Algorithm for Early Prediction of Vascular Compromise in 208 Microsurgical Flaps Using Tissue Oxygen Saturation Measurements,” Ann Plast Surg 62, 538–543 (2009).

13. A. Keller, “Noninvasive Tissue Oximetry for Flap Monitoring: An Initial Study,” J reconstr Microsurg 23, 189–197 (2007).

14. A. A. M. A. Lindelauf, A. G. Saelmans, S. M. J. van Kuijk, R. R. W. J. van der Hulst, and R. M. Schols, “Near-Infrared Spectroscopy (NIRS) versus Hyperspectral Imaging (HSI) to Detect Flap Failure in Reconstructive Surgery: A Systematic Review,” Life 12, 65 (2022).

15. “T. Ox. Tissue Oximeter Operator’s Manual,” (n.d.).

16. L. Johannesson, E. Richards, V. Reddy, J. Walter, K. Olthoff, C. Quintini, A. Tzakis, N. Latif, P. Porrett, K. O’Neill, and G. Testa, “The First 5 Years of Uterus Transplant in the US: A Report From the United States Uterus Transplant Consortium,” JAMA Surg (2022).

17. A. Baños, M. Wolf, C. Grawe, M. Stahel, D. Haensse, D. Fink, and R. Hornung, “Frequency domain near-infrared spectroscopy of the uterine cervix during cervical ripening,” Lasers Surg Med 39, 641–646 (2007).

18. R. Hornung, S. Spichtig, A. Baños, M. Stahel, R. Zimmermann, and M. Wolf, “Frequency-domain near-infrared spectroscopy of the uterine cervix during regular pregnancies,” Lasers Med Sci 26, 205–212 (2011).

19. R. Hornung, T. H. Pham, K. A. Keefe, M. W. Berns, Y. Tadir, and B. J. Tromberg, “Quantitative near-infrared spectroscopy of cervical dysplasia in vivo,” Hum Reprod 14, 2908–2916 (1999).

20. B. Matzinger, M. Wolf, A. Baños, D. Fink, and R. Hornung, “Optical properties, physiologic parameters and tissue composition of the human uterine cervix as a function of hormonal status,” Lasers Med Sci 24, 561–566 (2009).

21. N. T. Clancy, S. Saso, D. Stoyanov, V. Sauvage, D. J. Corless, M. Boyd, D. E. Noakes, M.-Y. Thum, S. Ghaem-Maghami, J. R. Smith, and D. S. Elson, “Multispectral imaging of organ viability during uterine transplantation surgery in rabbits and sheep,” J Biomed Opt 21, 106006 (2016).

22. S. Noori, B. Drabu, M. McCoy, and K. Sekar, “Non-invasive measurement of local tissue perfusion and its correlation with hemodynamic indices during the early postnatal period in term neonates,” J Perinatol 31, 785–788 (2011).

23. Y. Kagaya and S. Miyamoto, “A systematic review of near-infrared spectroscopy in flap monitoring: Current basic and clinical evidence and prospects,” J Plast Reconst Aesthet Surg 71, 246–257 (2018).

